# Evaluating the neuroprotective impact of senolytic drugs on human vision

**DOI:** 10.1101/2020.07.27.20163097

**Authors:** Nevin W. El-Nimri, Spencer M. Moore, Linda M. Zangwill, James A. Proudfoot, Robert N. Weinreb, Dorota Skowronska-Krawczyk, Sally L. Baxter

**Author notes:** Correspondence to: Sally L. Baxter, MD, MSc, Shiley Eye Institute, University of California, San Diego 9415 Campus Point Drive, MC 0946, La Jolla, CA 92093, Dorota Skowronska-Krawczyk, PhD, Department of Physiology and Biophysics, Department of Ophthalmology, Center for Translational Vision Research University of California Irvine, 837 Health Sciences Rd., Irvine, CA 92617. These authors contributed equally and should be considered first co-authors (Nevin W. El-Nimri, Spencer M. Moore). These authors contributed equally as co-corresponding authors (Dorota Skowronska-Krawczyk, Sally L. Baxter).

## Abstract

Glaucoma, a chronic neurodegenerative disease of retinal ganglion cells (RGCs), is a leading cause of irreversible blindness worldwide. Its management currently focuses on lowering intraocular pressure to slow disease progression. However, disease-modifying, neuroprotective treatments for glaucoma remain a major unmet need. Recently, senescent cells have been observed in glaucomatous eyes, exposing a potential pathway for alternative glaucoma therapies. Prior studies demonstrated that targeting senescent RGCs for removal (i.e., a senolytic approach) protected healthy RGCs and preserved visual function in a mouse ocular hypertension model. However, the effects of senolytic drugs on vision in human patients are unknown. Here, we used existing clinical data to conduct a retrospective cohort study in 28 human glaucoma patients who had been exposed to senolytics. Senolytic exposure was not associated with decreased visual acuity, elevated intraocular pressure, or documentation of senolytic-related adverse ocular effects by treating ophthalmologists. Additionally, patients exposed to senolytics (n=9) did not exhibit faster progression of glaucomatous visual field damage compared to matched glaucoma patients (n=26) without senolytic exposure. These results suggest that senolytic drugs do not carry significant ocular toxicity and provide further support for additional evaluation of the potential neuroprotective effects of senolytics on glaucoma and other neurodegenerative diseases.

## INTRODUCTION

Glaucoma is a chronic, progressive neurodegenerative disease of retinal ganglion cells (RGCs) whose axons form the optic nerve. Glaucoma is a leading cause of irreversible blindness worldwide and is expected to affect approximately 80 million people by 2020^1^. Currently, glaucoma management involves using topical medications and/or surgical procedures that lower intraocular pressure (IOP). Continued disease progression in patients with seemingly controlled IOP highlights the need to investigate other pathogenic mechanisms and potential alternative therapeutic targets^2^.

Based on neuropathological similarities of glaucoma with other age-related neurodegenerative diseases such as Alzheimer’s and the involvement of the ubiquitin-proteasome and chaperone systems, Caprioli and colleagues hypothesized a cellular senescence contribution to glaucoma pathogenesis^3^. Preclinical evidence has supported the cellular senescence hypothesis as a contributor to glaucoma pathogenesis. Homozygosity of glaucoma-risk alleles in *SIX6* in a glaucoma mouse model led to RGC senescence through *p16INK4a* overexpression^4^. Other data also suggest that TANK-binding protein 1 (TBK1), a known regulator of neuroinflammation, immunity, and autophagy, may respond to increased IOP and act downstream to regulate p16INK4a expression and RGC senescence^5^. Senescent cells secrete a plethora of molecules known as senescence associated secretory proteins (SASP), which affect surrounding cells by inducing either apoptosis or senescence, thus propagating the phenotype^6^. Since senescent cells are resistant to apoptosis themselves, they impact the tissue even after the initial stressor is removed. There are several senolytic drugs that are able to specifically target senescent cells to overcome the apoptosis block to remove them,^7,8^ presenting an attractive hypothesis for potential treatment of glaucoma. Indeed, our recent study has shown that targeting senescent RGCs in a mouse model of glaucoma using the senolytic drug dasatinib protected the remaining RGCs and visual function from glaucomatous injury^9^. These data are also supported by evidence from human studies, as a bioinformatics analysis of genes associated with primary open angle glaucoma suggested senescence as a key factor in pathogenesis^10^.

Little is known about the neuroprotective effects or safety of senolytic drugs on vision in human patients. Clinical management of glaucoma involves acquisition of extensive longitudinal data including visual acuity, IOP, visual field sensitivity and retinal nerve fiber thickness. Compared to other neurodegenerative diseases that often lack objective standardized metrics of clinical progression, some of these ophthalmic data are readily available and amenable to investigations of novel therapeutics, including senolytic drugs. To this end, we performed a retrospective analysis of existing clinical data to evaluate the effect of senolytics on vision and glaucoma progression. For the current study, we queried the electronic health record (EHR) system of a large academic medical center to identify glaucoma and glaucoma suspect patients exposed to at least one senolytic drug and conducted several analyses of visual data.

## RESULTS

### Patient Selection

Of 31,686 patients with glaucoma, glaucoma suspect, or ocular hypertension in our EHR clinical data warehouse, 111 unique patients (0.35%) were identified who had been exposed to a senolytic drug. Of these, 74 patients had ophthalmic data available (Figure 1).

**Figure 1.**
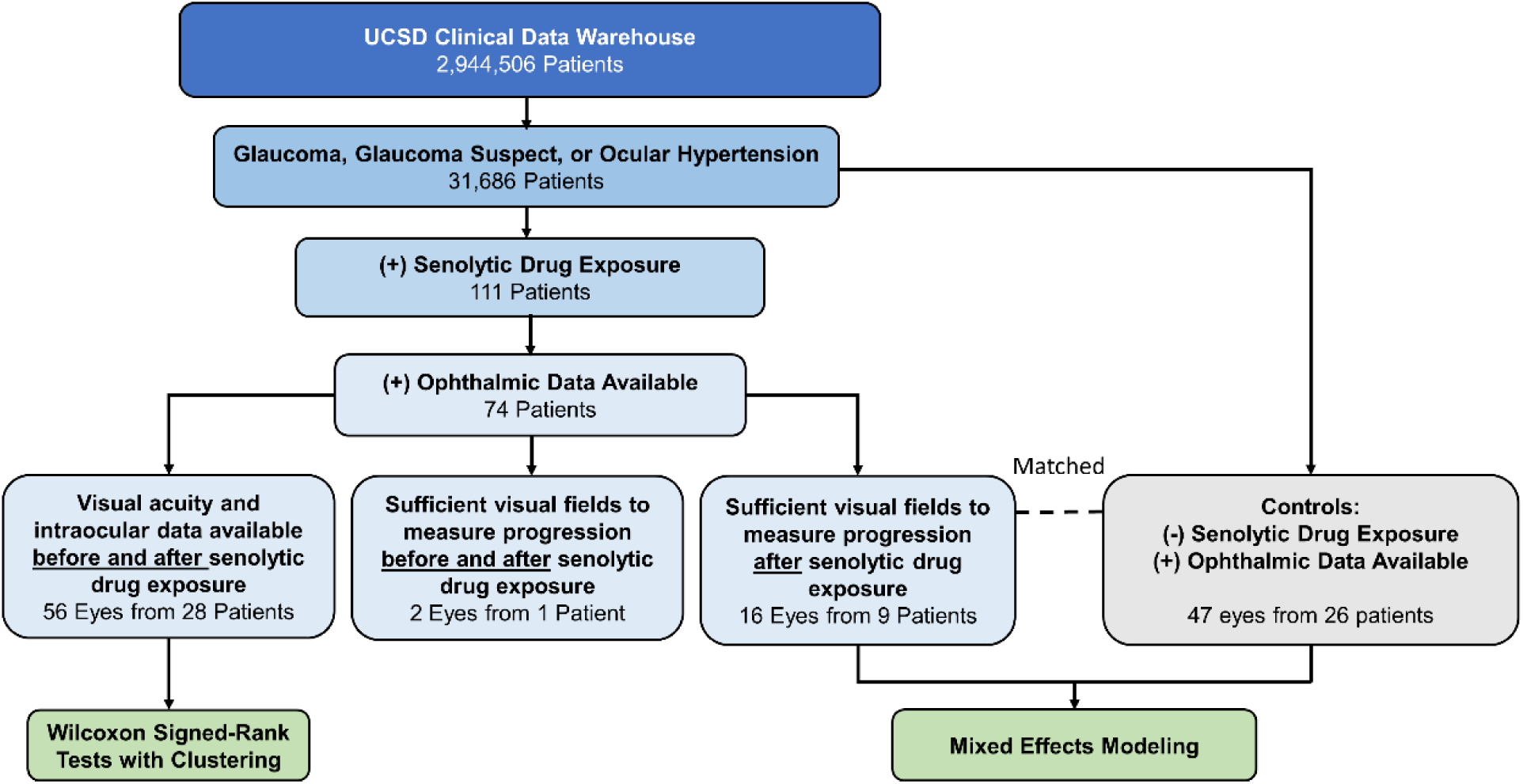
Overall workflow for evaluation of senolytic drug exposure on visual outcomes in glaucoma and glaucoma suspect patients. The University of California San Diego (UCSD) electronic health record clinical data warehouse was queried from 1/1/2005-3/16/2020. Visual field progression calculations were based on mean deviation values, and we required at least 3 reliable fields over minimum 1-year follow-up (reliability defined by <30% false positives, false negatives, and fixation losses). Controls were matched by age, sex, race, and baseline mean deviation.

### Senolytic Drug Exposures

Of the original *n*=74 cases included in the analysis, *n*=7 patients had been prescribed >1 senolytic drug (5 dasatinib-imatinib, 1 imatinib-nilotinib, 1 dasatinib-nilotinib). The most common senolytics were tocilizumab (n=20) and imatinib (n=19). Senolytic drug names, dosing, and numbers of patients are depicted in Figure 2. These drugs were most commonly prescribed for hematologic malignancies (such as chronic myeloid leukemia, acute myeloid leukemia, and acute lymphoblastic leukemia) and rheumatologic disorders. Other age-related or neurodegenerative diagnoses among these patients included age-related macular degeneration (*n*=3), cognitive impairment (*n*=2), and multiple sclerosis (*n*=1). In narrative clinical notes, no ocular adverse events were documented that were attributed to senolytic drug exposure.

**Figure 2.**
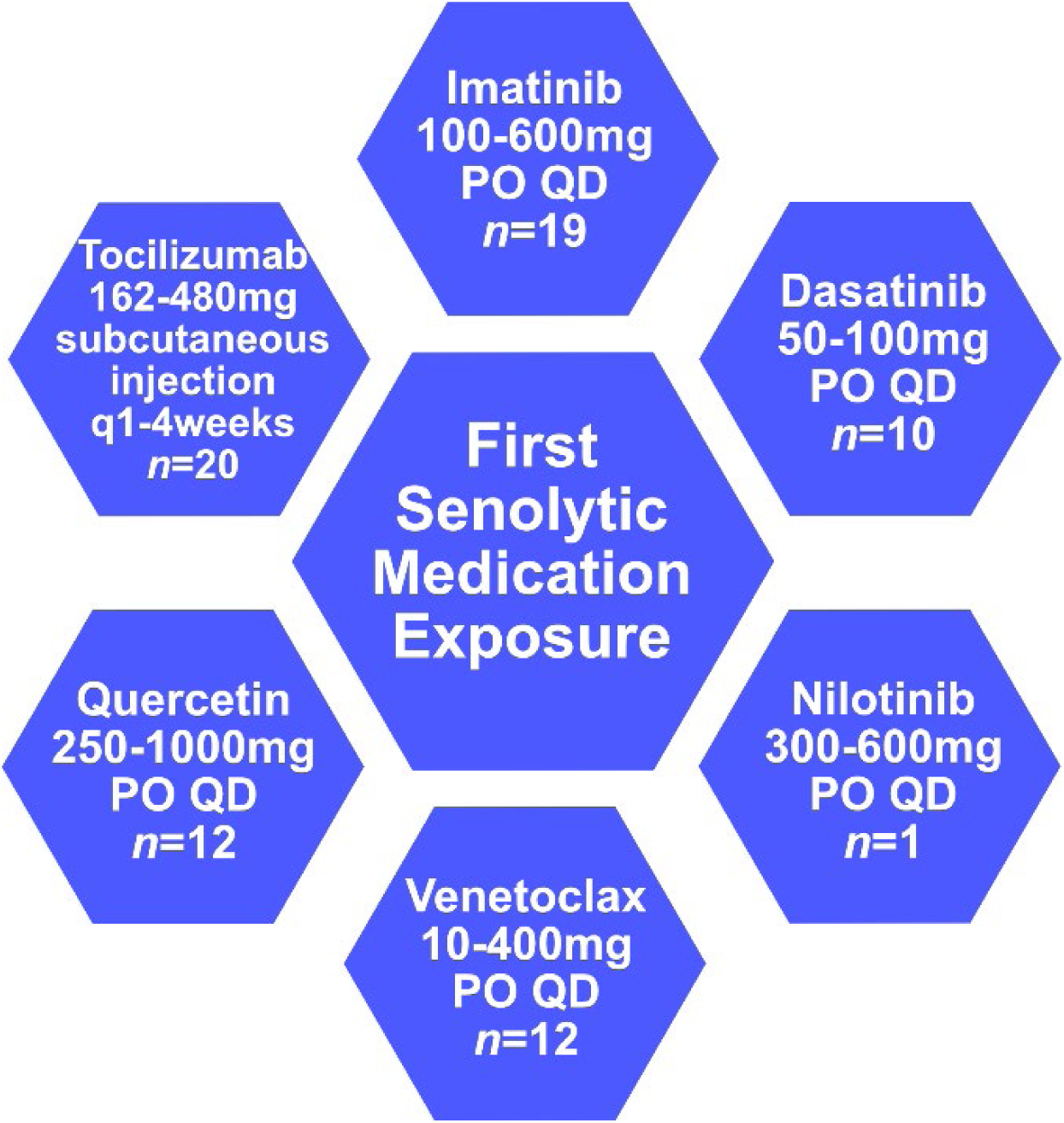
Generic names and dosage ranges of the first senolytic exposure in patients with glaucoma, glaucoma suspect, or ocular hypertension. PO= by mouth, QD= every day.

### Effects on Visual Acuity and Intraocular Pressure

For 56 eyes from 28 patients, sufficient data were available to examine visual acuity and intraocular pressure before and after senolytic medication exposure. The mean LogMAR visual acuity before senolytic medication exposure was 0.23 (equivalent to Snellen visual acuity of 20/34), which was not significantly different from the mean post-exposure logMAR visual acuity of 0.29 (Snellen 20/39) (p=0.28, see Figure 3A). Similarly, mean IOP was stable before and after senolytic medication exposure (17.3 vs. 16.9, p=0.34, Figure 3B). For these patients, the mean (standard deviation, SD) number of IOP-lowering medications during the follow-up period was 1 (1.1). Only two eyes from one patient had undergone any glaucoma laser procedure (argon laser trabeculoplasty), but this procedure pre-dated senolytic medication exposure by 8 years. Similarly for glaucoma surgeries, there were five eyes that had undergone trabeculectomies, and two eyes that had undergone glaucoma drainage device implantation, but in all cases, the glaucoma surgeries had been performed *before* senolytic medication exposure, ranging from 3 months to 13 years prior. Therefore, IOP stability after senolytic medication exposure was not confounded by glaucoma treatment.

**Figure 3.**
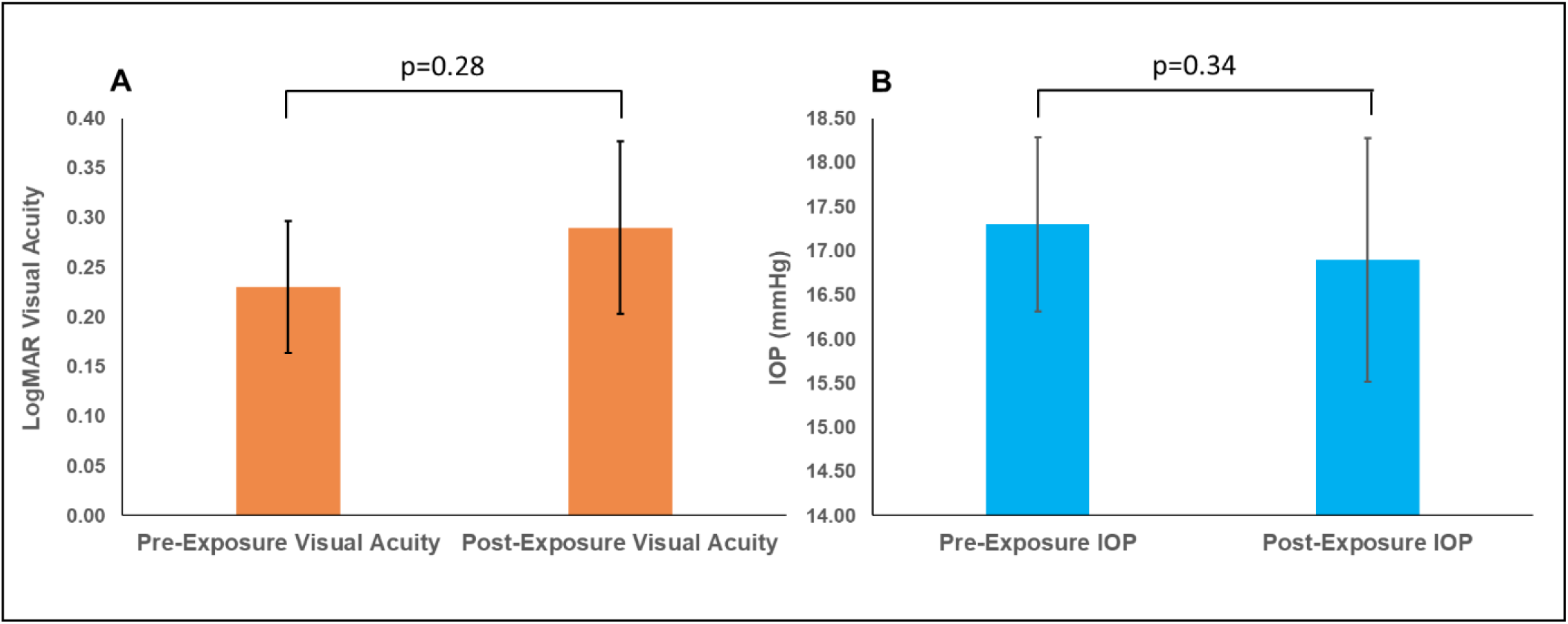
Effects of senolytics on visual acuity and intraocular pressure. Senolytic drug exposure was not associated with significant changes in visual acuity (panel A) or intraocular pressure (IOP) (panel B). Error bars denote standard error.

### Effects on Glaucoma Progression

To measure glaucoma progression, we analyzed trends of mean deviation (MD) values on visual fields. MD values are generally considered summary measures of disease severity, with more negative numbers indicating more glaucomatous damage and loss of sensitivity to light. Only one patient had sufficient (i.e., at least 3) visual fields to compare progression before and after senolytic drug exposure. This was a black male with primary open-angle glaucoma who was 73 years old at his first visual field (“baseline”). He was 76 years old when he was prescribed quercetin for inflammatory prostate disease. His last visual field before quercetin exposure was 6 months prior to taking the medication (“pre-exposure”). In his right eye, MD of the visual field was -3.81 dB at baseline, -3.57 dB pre-exposure, -1.75 dB one year after exposure, and -2.93 dB at his last visual field in that eye (five years after exposure). In his left eye, MD was -3.37 dB at baseline, -1.77 dB pre-exposure, -0.98 dB one year after exposure, and -2.16 dB at his last visual field in that eye (three years after exposure). The MD values of both eyes after quercetin exposure were better than pre-exposure, even with 3-5 years of follow-up. Formal statistical testing was not performed given the limited amount of data from a single patient.

We then compared visual field progression of 16 eyes of 9 individuals who had been exposed to a senolytic drug and had at least 3 reliable visual fields after exposure with a group of age, race, sex, and treatment matched patients (47 eyes of 26 individuals) without any history of senolytic drug exposure. There were no significant differences in the distributions of age, sex, or race between the two groups (Table 1). Similarly, although the mean (SD) baseline MD of - 8.02 [8.23] dB among eyes of exposed patients was lower than the mean (SD) baseline MD of - 5.60 (5.66) dB among eyes from non-exposed patients, this difference was not statistically significant given variation within each group (p=0.29). At baseline, there were no significant differences in number of IOP-lowering medications (mean of 0.59 in eyes of exposed patients, mean 0.47 in eyes of non-exposed patients, p=0.62). Similarly, there were no significant differences in number of glaucoma surgeries between the two groups (mean 0.12 in exposed patients, mean 0.22 in non-exposed patients, p=0.48). Patients without senolytic exposures had significantly longer mean follow-up times (mean [SD] of 10.9 [5.9] years, compared with 2.97 [1.9] years for exposed patients, p<0.001). The mixed effects model accounted for this by including length of follow-up and related interaction terms as covariates.

**Table 1.**
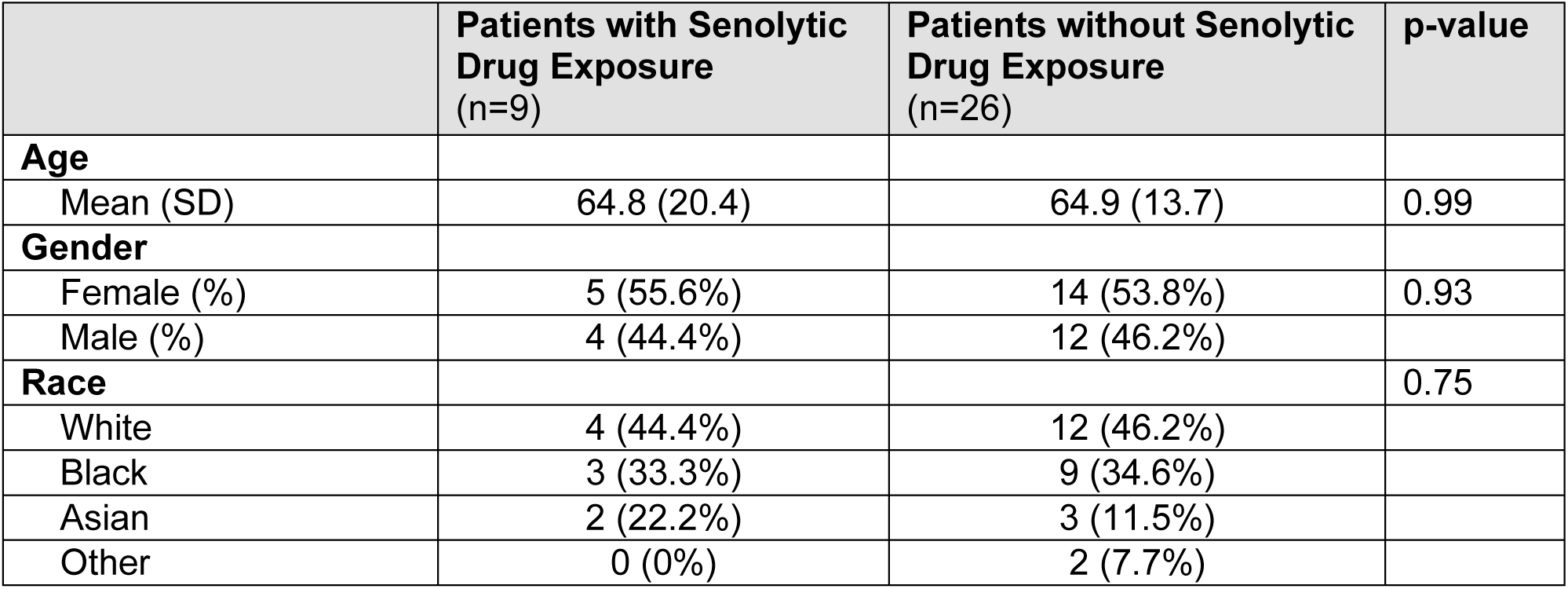
Demographics and clinical characteristics of patients with glaucoma, glaucoma suspect, or ocular hypertension and history of senolytic drug exposure and matched patients without senolytic drug exposure.

|  | **Patients with Senolytic Drug Exposure**<br>(n=9) | **Patients without Senolytic Drug Exposure**<br>(n=26) | **p-value** |
| --- | --- | --- | --- |
| **Age** |  |  |  |
| Mean (SD) | 64.8 (20.4) | 64.9 (13.7) | 0.99 |
| **Gender** |  |  |  |
| Female (%) | 5 (55.6%) | 14 (53.8%) | 0.93 |
| Male (%) | 4 (44.4%) | 12 (46.2%) |  |
| **Race** |  |  | 0.75 |
| White | 4 (44.4%) | 12 (46.2%) |  |
| Black | 3 (33.3%) | 9 (34.6%) |  |
| Asian | 2 (22.2%) | 3 (11.5%) |  |
| Other | 0 (0%) | 2 (7.7%) |  |

A mixed effects model of longitudinal changes in visual field MD demonstrated that senolytic drug exposure did not have a statistically significant effect on glaucoma progression (p=0.21). Figure 4 illustrates the strength of associations between covariates (y-axis) with changes in visual field MD (i.e., visual field deterioration/disease progression) based on coefficient estimates (x-axis). Covariates with coefficient estimates with 95% confidence intervals not crossing zero are significantly associated with changes in visual field MD. Not surprisingly, covariates significantly associated with accelerated visual field progression included more advanced baseline age, baseline MD, and the interaction between baseline MD and years of follow-up. Glaucoma treatment with medications, lasers, or surgeries also significantly influenced visual fields when monitored longitudinally, also unsurprising. Based on this model, senolytic drugs did not have any significant adverse effects on modulating visual field MD; i.e., they did not accelerate visual field decline.

**Figure 4.**
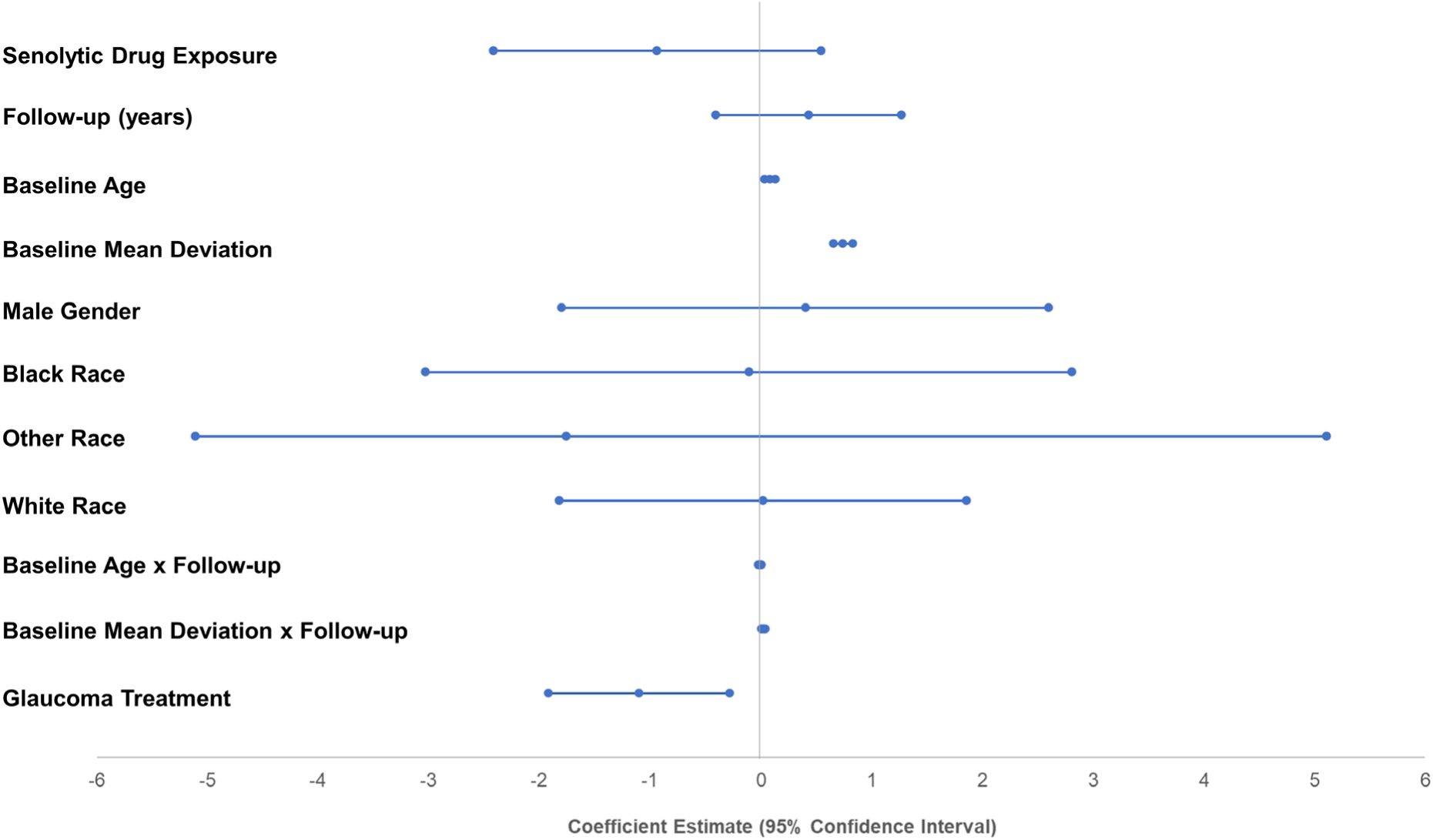
Mixed effects model of glaucoma progression based on changes in visual field mean deviation (MD). This plot depicts coefficient estimates and 95% confidence intervals for covariates in the model. Eyes and patients were included as random effects to account for within-subject correlation. Senolytic drug exposure (top-most interval) was not significantly associated with glaucoma progression as measured by changes in visual field MD (p=0.21).

## DISCUSSION

In this study, analysis of available data generated during routine clinical care did not identify evidence of ocular adverse effects associated with senolytic drug exposure. Specifically, these drugs were not associated with decreased visual acuity, elevated intraocular pressure, or accelerated visual field progression, even when accounting for other factors such as age, sex, race, and glaucoma treatment. These findings provide reassuring evidence regarding the safety profile of senolytic drugs for the visual system.

Age is a well-recognized risk factor for glaucoma development and likely contributes to optic nerve head vulnerability and lessened ability to resist damage from elevated IOP over time^11-17^. Previous studies have shown that aged/senescent cells in the outflow pathway and among RGCs are found in glaucoma and experimental ocular hypertension^3,4,9^. Therefore, there has been concern that senolytic drugs taken by older glaucoma patients could jeopardize their limited remaining RGCs, potentially advancing visual impairment. Fortunately, our results provided evidence for a favorable safety profile based on clinical data, although more advanced imaging technologies or postmortem analysis are needed to determine effects of senolytic exposure on RGC density and viability.

Our results also have potential implications for other neurodegenerative diseases outside of glaucoma. There is considerable evidence that supports the notion of a common pathogenic mechanism between glaucoma and other neurodegenerative diseases such as Alzheimers and Parkinsons^18^, including caspase protease activation^19^, microglial-mediated neuroinflammation^20^, and platelet-generated amyloid peptides^21^. With shared pathways, strategies developed to treat glaucoma could also be potentially applied to other neurodegenerative disorders^19^. However, an important challenge in monitoring other neurodegenerative diseases is that formal neurocognitive testing is rarely performed outside research settings. In contrast, imaging and visual field tests are frequently performed by eyecare professionals as part of their routine clinical care. With proper validation, objective visual data, such as visual acuity and visual fields, could potentially serve as a biomarker or proxy for monitoring progression and treatment response of other neurodegenerative conditions, particularly since age-related vision disorders/changes often precede other systemic conditions^22-26^. Given the ties between glaucoma and other neurodegenerative diseases, as well as our findings of vision-related safety of senolytic drugs, glaucoma offers an ideal opportunity to test the potential neuroprotective effects of using drugs targeting senescent cells.

Several items should be considered when interpreting our results. The size of our patient cohorts was a key limitation of this study that constrained generalizability to larger populations. Of the 111 glaucoma patients with senolytic exposure, 28 patients had available visual acuity and IOP data before and after exposure, and only 9 patients had sufficient visual fields to measure progression post-exposure. These relatively small number may be related to the senolytics being relatively recent therapeutic options. Imatinib, the first to be approved, was cleared by the United States Food and Drug Administration (FDA) in 2001. This was also evidenced by the fact that our exposed patients had relatively short follow-up periods with a mean of ∼3 years. Future studies combining data from other centers and following additional patients longitudinally for longer periods of time are likely to provide additional insights into the effects of senolytics on vision and glaucoma. Further, the small sample size led to aggregation of senolytic-exposed patients into a single group for analysis, which may represent an oversimplification, as tyrosine kinase inhibitors such as imatinib may exert senolytic effects by a distinct mechanism from other drugs like tocilizumab, an anti-IL6 receptor monoclonal antibody.

Patients in our cohort were prescribed senolytics for a variety of indications, such as treatment of hematologic malignancies, autoimmune or rheumatic diseases, or as dietary supplementation. Dosing for these drugs’ primary indication is likely to exceed that required for senolytic effects in the central nervous system and eyes. Rocha et al. used 5mg/kg dasatinib in the mouse model to achieve senolytic effects in RGCs^9^, which converts to approximately 0.4mg/kg in human equivalent dosing. When accounting for species differences in drug metabolism^27^, this dosage is less than the approximately ∼1mg/kg or a higher dose prescribed to many patients in our study. A theoretical senolytic regimen to slow glaucoma progression may also resemble treatment of chronic illness (i.e., chronic long-term treatment) rather than the shorter, high-dose treatments for hematologic malignancy or rheumatic disease experienced by the patients in our study. Even for cancer treatments, optimal senolytic dosing has not yet reached consensus^28^, so this remains an ongoing area of investigation. Another limitation to the generalizability of our data is patients’ general health concerns, as those with senolytic exposure had significant medical comorbidities such as hematologic malignancy and rheumatic disease. Finally, as this is an observational study, no conclusions regarding causation can be drawn.

In conclusion, we have leveraged existing clinical data to investigate the safety of senolytics on vision-related phenotypes in glaucoma patients. The present study suggests no association between senolytic drug exposure and adverse ocular effects or alteration of the rate of glaucoma progression. Further studies with larger patient cohorts will be needed in conjunction with ongoing work in preclinical animal models. Moreover, insights from glaucoma patients could motivate additional trials of senolytic drugs in other age-related neurodegenerative diseases.

## METHODS

### Study Design and Population

This was a retrospective cohort study examining medical record data for adult patients with glaucoma, glaucoma suspect, or ocular hypertension receiving ophthalmic care at the University of California San Diego (UCSD), an academic medical center in La Jolla, CA, between January 2005 and March 2020. This study was approved by the UCSD Institutional Review Board (IRB) and adhered to the tenets of the Declaration of Helsinki. Waiver of informed consent was also granted by the UCSD IRB.

### Characterizing Senolytic Medication Exposure and Visual Outcomes

For overall study workflow, see Fig. 1. The exposure of interest was senolytic drug intake. We identified patients by querying the UCSD clinical data warehouse using a tool embedded in the enterprise electronic health record (EHR) system (Epic SlicerDicer, Epic Systems, Verona, WI). Using International Classification of Disease (ICD) codes, we searched for patients with a diagnosis of glaucoma, glaucoma suspect, or ocular hypertension in their chief complaint, medical history, problem list, or billing diagnoses. Among these 32,414 patients, we identified 111 patients with orders for the following senolytic drugs: imatinib, dasatinib, venetoclax, nilotinib, tocilizumab, and quercetin.

Among this group of 111 patients, 74 had received ophthalmic care at our institution. For these, we recorded duration, dose, and frequency of all senolytic drugs. We also recorded the indication for the senolytic drug as recorded in the EHR by the ordering physician. We recorded visual acuity and intraocular pressure both before and after senolytic medication exposure, requiring at least three follow-up visits after medication exposure. We also noted whether any adverse ocular effects had been recorded in the patient’s medical record that were documented by treating physicians as attributable to senolytic medication exposure. We extracted information on glaucoma medications, lasers, and surgeries. For patients with available visual field data, we required at least 3 Humphrey 24-2 visual fields (Zeiss, Oberkochen, Germany) of adequate reliability (defined as <30% false positives, false negatives, and fixation losses) to calculate glaucoma progression based on changes in mean deviation (MD).

### Statistical Analyses

We first evaluated whether there were any significant differences in visual acuity or intraocular pressure associated with senolytic exposure. To do this, we converted Snellen visual acuity measurements to logMAR equivalents using previously published methods^29^. Wilcoxon signed-rank tests with clustering were performed using the *clusrank* package in R^30^, with each eye’s average pre- and post-exposure measurements serving as a matched pair and each patient representing a cluster to account for correlation between eyes. Next, to evaluate the effects of senolytic exposure on glaucoma progression, we intended to compare visual field progression before and after senolytic medication exposure. However, only a single patient had at least three reliable visual fields both before and after medication exposure. Therefore, we decided to compare post-exposure visual fields for patients with senolytic exposure to visual fields from matched patients without any history of senolytic exposure. These patients were matched by age, gender, race, baseline MD, and glaucoma treatment (e.g. number of IOP-lowering medication and number of glaucoma procedures) at approximately a 3:1 ratio overall, given that higher ratios generally do not provide additional power^31^. To compare visual field progression, we generated a multivariable mixed effects model of change in MD with history of senolytic drug exposure, length of follow-up, baseline age (defined as age at first visual field), baseline MD (defined as MD at first visual field), sex, race, and glaucoma treatment designated as co-variates. We included interaction terms between baseline age and follow-up as well as between baseline MD and follow-up. Eyes and patients were included as random effects to account for within-subject correlation. Statistical significance was defined as p<0.05 for all hypothesis testing and modeling. All statistical analyses were performed in R version 3.5.1. De-identified data and analysis notebooks are contained in an open-source repository^32^.

## Data Availability

All data referred to in the manuscript came from patients medical charts and secure healthcare software (EPIC). The processed data is saved in a secure university drive with protected password.

## Abbreviations list

EHR: Electronic Health Record
LogMAR: Logarithm of the Minimum Angle of Resolution
MD: Mean Deviation
PO: By Mouth
QD: Everyday
RGC: Retinal Ganglion Cell
SD: Standard Deviation

## Author Contributions statement

NEN: Contributed to the design of the study, data collection, data interpretation, and manuscript writing/revision.

SMM: Contributed to the design of the study, data collection, data interpretation, and manuscript writing/revision.

LMZ: Contributed to data interpretation and substantial manuscript revision.

JAP: Contributed to data analysis, data interpretation, and manuscript revision.

RNW: Contributed to data interpretation and substantial manuscript revision.

DSK: Contributed to the design of the study, data interpretation, and substantial manuscript revision.

SLB: Contributed to the design of the study, data analysis, data interpretation, and manuscript writing/revision.

## Funding/Support

The study was supported by grants from the National Institutes of Health (T15LM011271, P30EY022589, EY027011, EY11008, EY19869, EY14267, EY027510, EY026574, EY027945, EY029058) and by RPB Unrestricted Grant to Shiley Eye Institute and Gavin Herbert Eye Institute. Work in the DSK laboratory was additionally supported by RPB Special Scholar Award and Glaucoma Research Foundation Shaffer Award.

### Financial Disclosures

NEN: No financial disclosures to report.

SMM: No financial disclosures to report.

LMZ: F: National Eye Institute, Carl Zeiss Meditec Inc., Heidelberg Engineering GmbH, Optovue Inc., Topcon Medical Systems Inc. R: Heidelberg Engineering, P: Zeiss Meditec.

JAP: No financial disclosures to report.

RNW: Research support–Carl Zeiss Meditec, Centervue, Heidelberg Engineering, Konan, National Eye Institute, Optovue; Consultant–Aerie Pharmaceuticals, Allergan, Bausch & Lomb, Eyenovia.

DSK: No financial disclosures to report.

SLB: SLB is supported by the National Institutes of Health (T15LM011271).

